# Effect of an Artificial Intelligence Chest X-Ray Disease Prediction System on the Radiological Education of Medical Students: A Pilot Study

**DOI:** 10.1101/2022.07.11.22277278

**Authors:** Lan Dao, Sabrina Sofia Harmouch, Anne Chin, Tien Dao, Zhe Thian, Carl Chartrand-Lefebvre, Joseph Paul Cohen

**Author notes:** Corresponding author: Lan Dao, University of Montreal, 2900 Boulevard Edouard-Montpetit, QC H3T 1J4, Montreal, Quebec, Canada. This study has been approved by the Institutional Review Board of the University of Montreal.

## Abstract

**BACKGROUND:** We aimed to evaluate the feasibility of implementing Chester, a novel web-based chest X-ray (CXR) interpretation artificial intelligence (AI) tool, in the medical education curriculum and explore its effect on the diagnostic performance of undergraduate medical students.

**METHODS:** Third-year trainees were randomized in experimental (N=16) and control (N=16) groups and stratified for age, gender, confidence in CXR interpretation, and prior experience. Participants filled a pre-intervention survey, a test exam (Exam1), a final exam (Exam2), and a post-intervention survey. The experimental group was allowed to use Chester during Exam1 while the control group could not. All participants were forbidden from using any resources during Exam2. The diagnostic interpretation of a fellowship-trained chest radiologist was used as the standard of reference. Chester’s performance on Exam1 was 60%. A five-point Likert scale was used to assess students’ perceived confidence before/after the exams as well as Chester’s perceived usefulness.

**RESULTS:** Using a mixed model for repeated measures (MMRM), it was found that Chester did not have a statistically significant impact on the experimental group’s diagnostic performance nor confidence level when compared to the control group. The experimental group rated Chester’s usefulness at 3.7/5, its convenience at 4.25/5, and their likelihood to reuse it at 4.1/5.

**CONCLUSION:** Our experience highlights the interest of medical students in using AI tools as educational resources. While the results of the pilot project are inconclusive for now, they demonstrate proof of concept for a repeat experiment with a larger sample and establish a robust methodology to evaluate AI tools in radiological education. Finally, we believe that additional research should be focused on the applications of AI in medical education so students understand this new technology for themselves and given the growing trend of remote learning.

## 1. Introduction

In the last decades, artificial intelligence has demonstrated its value to improve efficiency and productivity in radiology (1–6). There is growing evidence that the training of post-graduate trainees and medical students should include understanding of artificial intelligence (AI) (7–10). While prior research has focused on the use of AI in precision medicine and its clinical applications, AI tools applications to medical education remain underexplored. Agent-assisted learning (software-augmented learning) has been proposed as a distance education tool to supplement class lectures and reform medical education curricula (11,12). To date, few AI tools have been proposed for education in radiology. To date, the Adaptive Radiology Interpretation and Education System (ARIES) is one of the few systems proposed for education in radiology, focusing on quantitative characterization of brain MRIs to support diagnosis (13,14). In this context, the Mila (Quebec Artificial Intelligence Institute) medical research group released Chester, a web-based, locally run system for diagnosing frontal chest X-rays in 2019 (15). The system comprises three sections: an input image of a patient’s chest X-ray; a disease prediction including continuous scales of 14 radiological findings from “healthy” to “at risk”; and a saliency map showing regions influencing the prediction (predictive image regions serving as prediction explanation) (16). This AI radiology assistant prototype is unique in its accessibility and ease of use, being a free and open tool easily installed as a desktop application, a web page, or on a cell phone, allowing any user to identify radiographic findings on chest X-ray images while preserving privacy (15,16). While code is delivered to a web browser, all processing occurs locally as patient data stays on the user’s machine (16). The system was evaluated on many external datasets and reported performance of the software ranges from 0.72 to 0.93 for different disease labels on chest X-ray images (17,18).

In this study, we aimed to fill the gap in the literature regarding AI tools and the feasibility of its use in the medical curriculum by designing a pilot randomized study to explore the impact of Chester on the diagnostic performance and confidence level of third-year medical students in frontal chest X-rays interpretation. We also aimed to explore the acceptability of the tool. We hypothesized that this educational intervention would increase medical trainees’ knowledge compared to the control cohort and would be a well-received tool among medical students.

## 2. Materials and Methods

### 2.1 Tool

Chester’s 14 findings encompass atelectasis, cardiomegaly, effusion, infiltration, mass, nodule, pneumonia, pneumothorax, consolidation, edema, emphysema, fibrosis, pleural thickening, and hernia (16). Chester’s diagnostic performance for Exam1 was exactly 60%, meaning that it correctly identified findings that were present with 60% accuracy. A finding was considered successfully identified when it was correctly shown on the saliency map. Indeed, for any high probability finding, the saliency map reveals the pixels Chester’s algorithm bases itself on to make a specific prediction when the user clicks on the “explain” button: for example, Figure 1 shows three high probability findings: pneumonia, infiltration, and lung opacity. Incorrectly identified findings were defined as Chester either omitting a finding that was present or falsely recognizing a finding that was not present.

**Figure 1:**
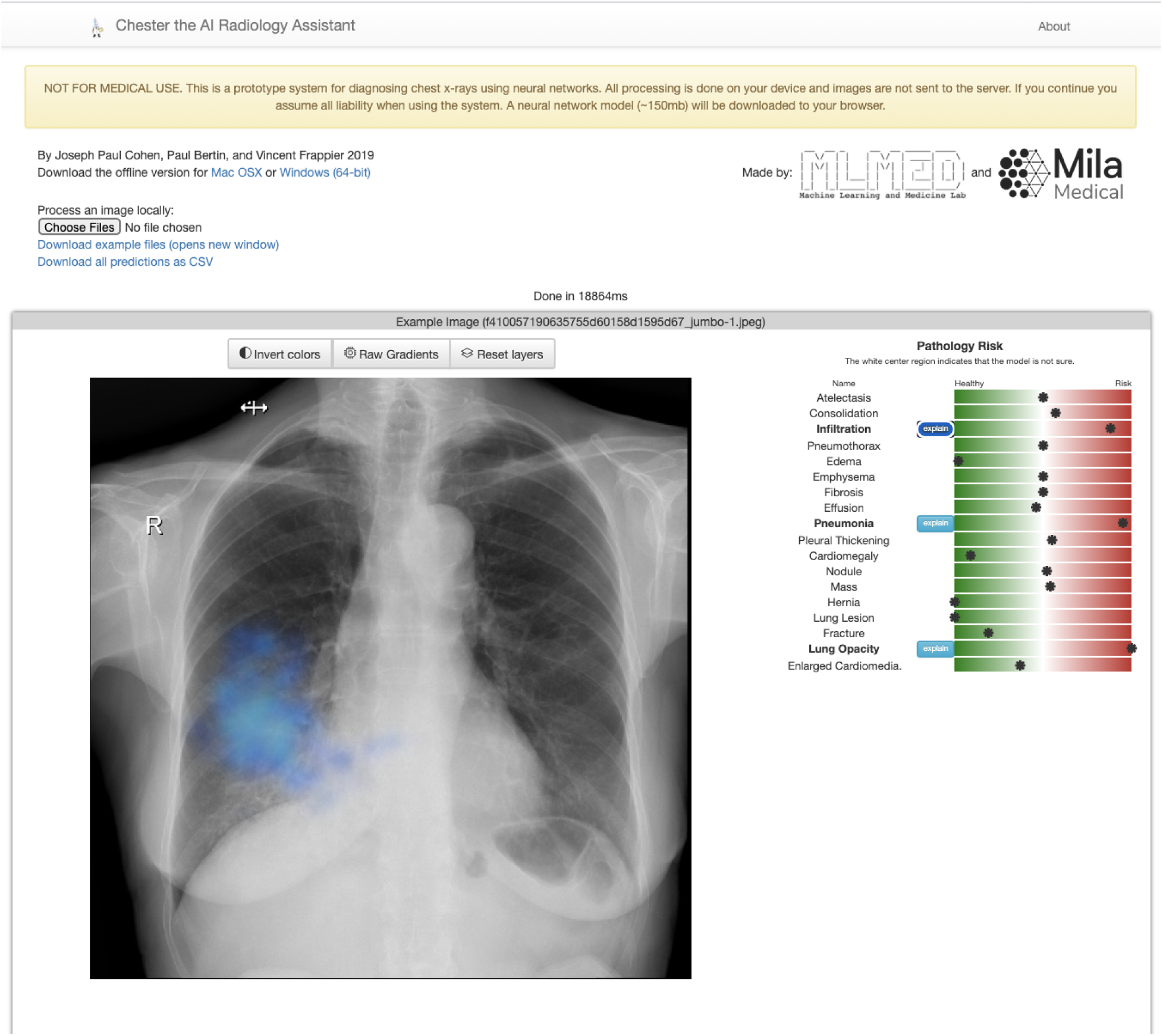
Interface of the artificial intelligence radiology assistant tool (Chester) displaying the saliency map feature for the finding “infiltration”

### 2.2 Study setting

This study was conducted remotely in collaboration with the University of Montreal’s Faculty of Medicine on a cohort of 32 third-year medical students. The study was held outside the context of a pre-existing course. All students, whether they agreed to participate in the study or not, had completed a respiratory medicine course as part of the curriculum during their second year of medical school. This course included a lecture by radiologists from the university’s Radiology Department comprising an introduction to chest X-ray interpretation and guidelines to identifying common radiological findings, including atelectasis, cardiomegaly, pleural effusion, lung mass, pulmonary nodule, lung infiltration, pneumonia, lung consolidation, pneumothorax, pulmonary edema, emphysema, idiopathic pulmonary fibrosis, pleural thickening, and diaphragmatic hernia.

During the study’s promotional campaign, students were introduced to the outline of the intervention and shown a link to the tool^1^ (Figure 1) as well as a video presentation explaining its features, overall functioning, limitations, and recommended use for medical students^2^ (Figure 2). In short, they were advised to use Chester during the practice test by processing the image of a given case in the platform, they were told that the system was imperfect, and they were encouraged to use their critical and clinical thinking skills as well as other resources if they saw fit. Participants were recruited via student groups’ mailing list and through posts on student Facebook groups in the weeks preceding the intervention.

**Figure 2:**
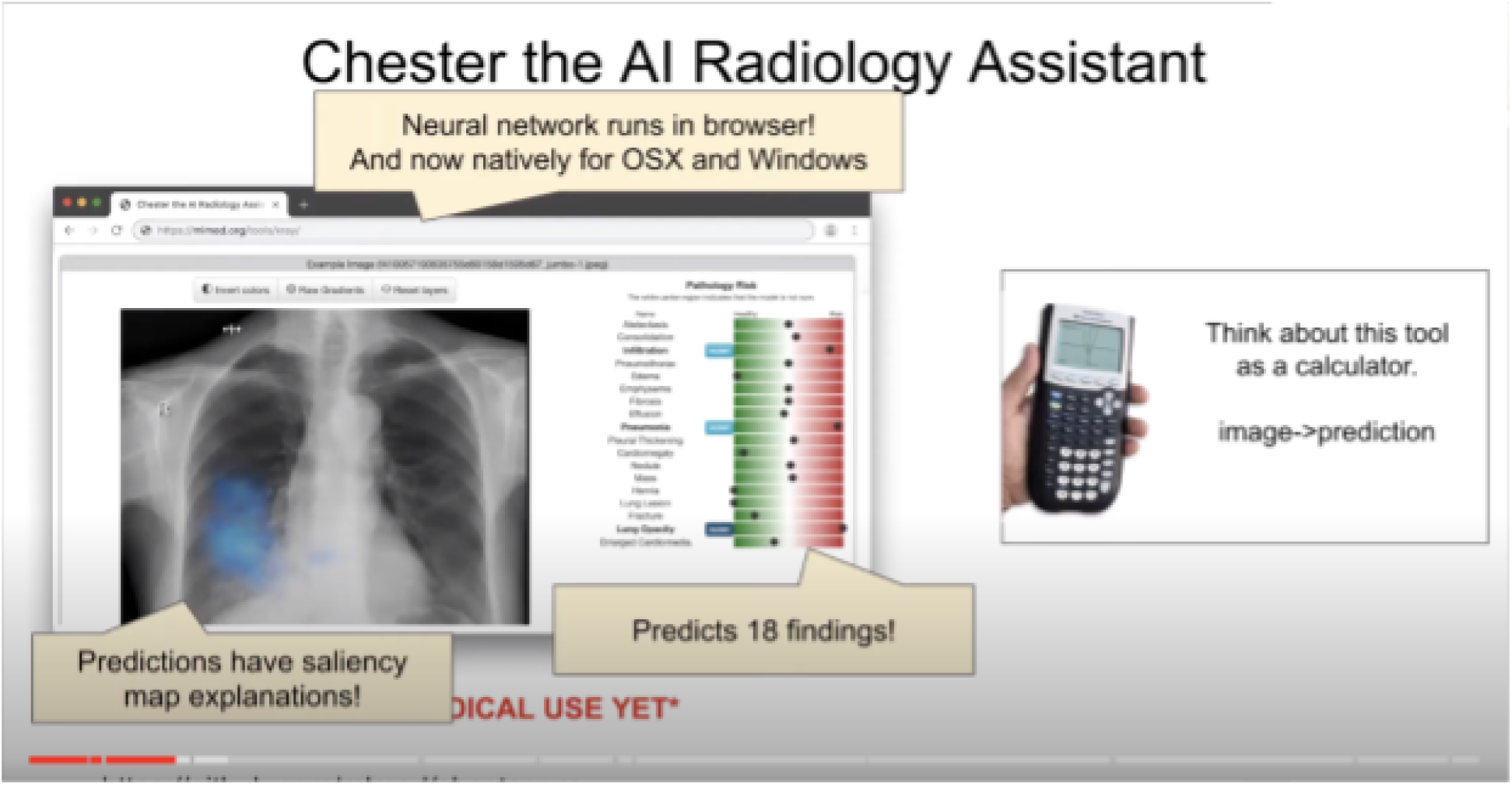
Video presentation sent to all students during the recruiting process and to participants in the intervention group (Chester) prior to the experiment

### 2.3 Study Design

The intervention comprised a pre-intervention survey, a test exam (Exam1), a final exam (Exam2), and a post-intervention survey. Conditions for eligibility included being a third-year medical student at the University of Montreal. Conditions for ineligibility included students who did not answer all 4 questionnaires of the intervention (Flowchart 1).

### 2.4 Pre-intervention survey

Students who were interested to participate to the study filled a pre-intervention survey (Supplementary Digital Content 1) collecting information such as participants’ year of study, their gender, their age, whether they had received education regarding chest X-ray interpretation or completed a clerkship rotation in a radiologic specialty, and their confidence levels in the interpretation of chest X-ray images on a 5-point Likert scale. Upon reception of the surveys, stratified randomization was performed in order to balance out prognostic variables.

### 2.5 Randomization

Participants were then randomized into two groups based on their gender, their age, whether they had received education regarding chest X-ray interpretation or completed a clerkship rotation in a radiologic specialty, and their confidence level in the interpretation of chest X-ray images on a 5-point Likert scale. Thus, there was an experimental group, which had access to the AI assistant tool during the practice test (Exam1), and a control group, which did not have access to the AI assistant tool during both Exams1 and Exam2. Subsequent to randomization, participants received an email containing their identification number with their group (experimental or control).

### 2.6 Practice test (Exam1)

Participants then received a link to the practice test Exam1 (Supplementary Digital Content 2 and Supplementary Digital Content 3). Since the study was conducted remotely, before accessing the questions in Exam1, participants were asked to sign a sworn statement stipulating that they would complete the questionnaire themselves, without help from other people.

Students completed an online multiple-choice practice test on Google Forms consisting of 15 frontal chest X-ray images accompanied by case notes (Figure 3): their task was to identify the main radiological finding in each case, which contained one or more of the 14 possible findings. The images were chosen from Radiopaedia^3^, a platform for radiology images made publicly available under a modified creative commons license. Both Exam1 and Exam2 (images and case notes) as well as their correction keys were approved by a fellowship-trained chest radiologist as appropriate for the level of the participants. Each correct case counted as 1 point, adding up to a total of 15 points. Lung infiltration, lung consolidation, and pneumonia were grouped as one entity, which meant that there were 12 answer choices on the test. Chester’s “lung opacity” and “lung lesion” findings were removed from the answer choices due to their lack of specificity.

**Figure 3:**
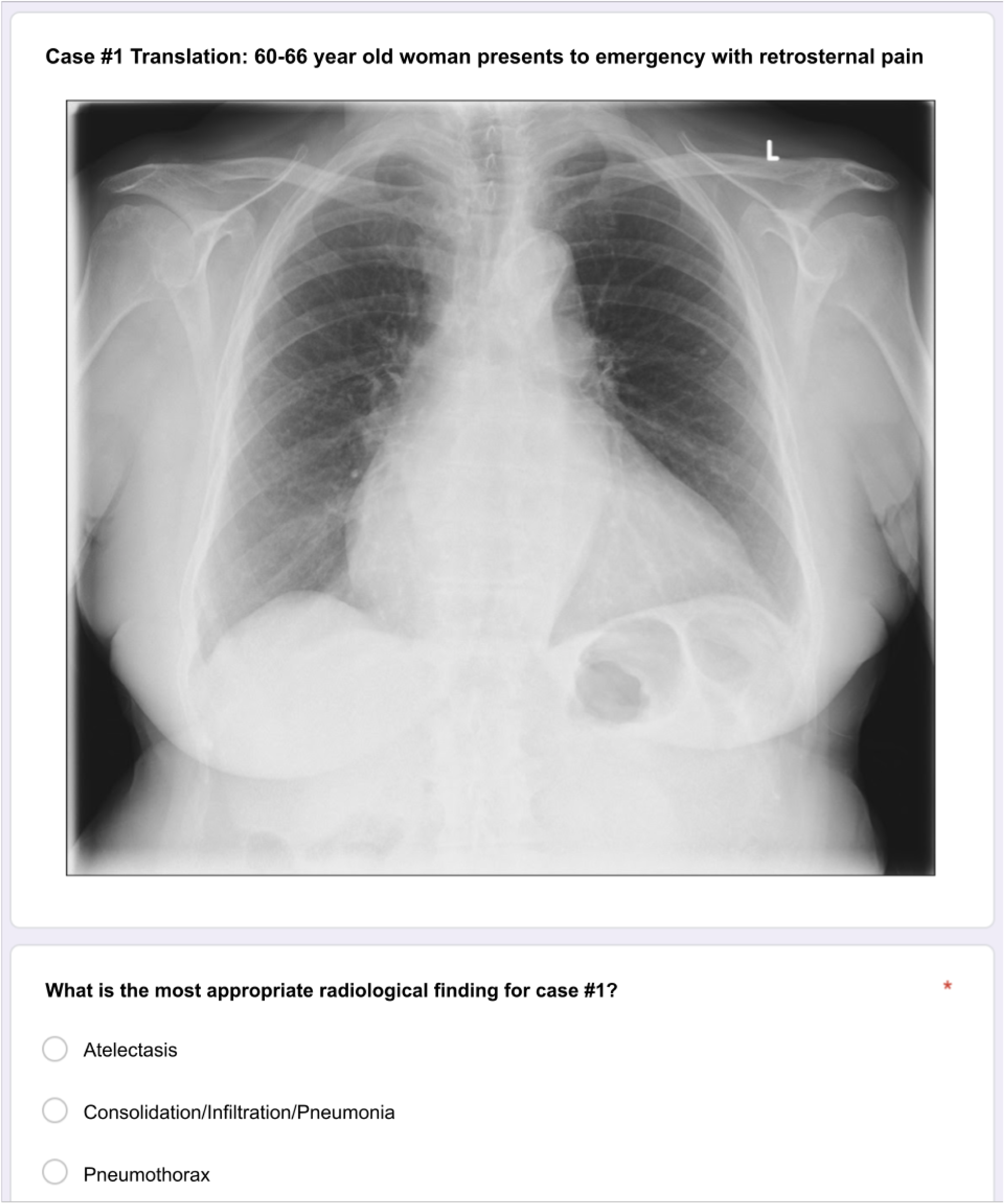
Example of a question with a chest X-ray image and accompanying case notes. Note this example is in English but the test was given in French. The example image is from the public Chest X-ray14 dataset.

During Exam1’s completion, the experimental group was given access to Chester through a URL as well as the presentation containing instructions on its usage, both of which were already linked in promotional posts featured on student groups. Participants in the control group, by contrast, had no access to Chester. Furthermore, both the experimental and control groups were given permission to use any available material, including online searches. There was no fixed time for the completion of Exam1. Following submission of the practice exam, participants received immediate retroaction (including any missed questions), correct answers and their explanations in the form of an answer key, and point values.

### 2.7 Final test (Exam2) and evaluation of the intervention

Subsequently, participants received a link to Exam2, the final test, which had the same format as Exam1 for consistency purposes (Supplementary Digital Content 4). Students were asked to identify themselves using their previously issued identification number and signed the same sworn statement as for Exam1. During this individual assessment period, no participant from the experimental nor the control group was permitted any material, including Chester and online searches. As for Exam1, there was no fixed time for the completion of Exam2. Following the submission of the final exam, participants also received immediate retroaction (including any missed questions), correct answers and their explanations in the form of an answer key, and point values.

### 2.8 Post-intervention survey

Finally, after receiving exam feedback, participants were given a link to an online post-intervention survey (Supplementary Digital Content 5). Using a 5-point Likert scale, participants from the experimental group were asked to assess (i) their level of confidence in their chest X-ray interpretation skills following the final exam, (ii) the perceived usefulness of the AI assistant tool, (iii) their willingness to use it if they were not prompted to do so, (iv) their level of trust in it, (v) how much they used it as opposed to other educational material, (vi) their perceived level of advantage (or disadvantage) compared to the control group, and (vii) how much they would recommend it to other medical students for learning. In comparison, students from the control group were asked to assess (i) their level of confidence in their chest X-ray interpretation skills following the final exam, (ii) the perceived usefulness of the AI assistant tool, (iii) their willingness to use it had they been given the opportunity to do so, and (iv) their perceived level of advantage (or disadvantage) compared to the experimental group. Participants from both groups were offered the opportunity to leave written comments at the end of the survey.

### 2.9 Statistical Analysis

Characteristics of both groups were analysed. Means and standard deviations were calculated for all continuous variables and proportions were reported for all categorical variables. A mixed model for repeated measures (MMRM) was used to analyze the data (19). This statistical model has been shown to be superior to repeated measures ANOVA in prior research because it underscores patterns of change after an intervention while accounting for individual differences. To analyze Chester’s impact on students’ diagnostic performance, parameters of the regression model were fit for *mx* + *b* against Exam1 versus Exam2, using the experimental group and the control group as independent covariates along with their interaction. To analyze Chester’s impact on students’ confidence, the same model was fit against the pre-intervention and post-intervention questionnaires with the same setup for independent covariates. All *p-values <* 0.05 were considered significant. All statistical analyses were performed using R version 4.0.3.

## 3. Results

32 third-year medical students from the University of Montreal participated in this randomized pilot study. The mean age was 24 years old. 56.3 % (18) of students were women and 43.8 % (14) of students were men. The median level of confidence in chest X-ray interpretation skills in the pre-intervention survey was 2 on a scale of 1 to 5, five being the most confident. Following randomization, all 32 students completed all four phases of the experiment. Baseline characteristics of each group are presented in Table 1. The de-identified responses to all four questionnaires (pre-test survey, Exam1, Exam2, post-test survey) are available in a public dataset. Results are graphically presented as least squares mean estimates along with their confidence intervals in Table 2 and Table 3, while a visualization of the MMRM estimates is plotted in Figure 4 and Figure 5.

**Table 1:** Baseline characteristics of participants involved in the study, including responses to pretest survey (level of confidence as well as completion of clerkship-level radiology workshop and extracurricular training in radiology)

| Characteristics |  | All participants | Control group (no Chester) | Intervention group (Chester) |
| --- | --- | --- | --- | --- |
| N (%) |  | 32 | 16 (50%) | 16 (50%) |
| Age in years | Mean (range) | 24.4<br>(21.0-38.0) | 24.8 (21.0-38.0) | 24.1 (21.0-29.0) |
|  | Median | 23.0 | 23.0 | 23.0 |
| Gender | Female n, (%) | 18 (56.2) | 9 (56.2) | 9 (56.2) |
|  | Male n, (%) | 14 (43.8) | 7 (43.8) | 7 (43.8) |
| Level of confidence in CXR interpretation (1 to 5, 5 being maximum confidence) | Mean (range) | 2.2 (1.0-3.0) | 2.3 (1.0-3.0) | 2.1 (1.0-3.0) |
|  | Median | 2.0 | 2.0 | 2.0 |
| Clerkship-level radiology workshop completed, n (%) | No | 25 (78.1) | 11 (68.8) | 14 (87.5) |
|  | Yes | 7 (21.9) | 5 (31.2) | 2 (12.5) |
| Extracurricular training in radiology completed, n (%) | No | 17 (53.1) | 8 (50.0) | 9 (56.2) |
|  | Yes | 15 (46.9) | 8 (50.0) | 7 (43.8) |

*CXR: chest X-ray*

**Table 2:** Participant’s diagnostic performance for the practice test (Exam1) and the final test (Exam2)

| Intervention | Group | Mean diagnostic performance | 95% Confidence interval |
| --- | --- | --- | --- |
| Exam1 | Control (Group B) | 69.17 | [ 62.69 ; 75.65 ] |
|  | Experimental (Group A) | 72.08 | [ 65.60 ; 78.56 ] |
| Exam2 | Control (Group B) | 78.33 | [ 71.85 ; 84.81 ] |
|  | Experimental (Group A) | 76.67 | [ 70.19 ; 83.15 ] |

**Table 3:** Participants’ confidence level in the pre-intervention and post-intervention questionnaires

| Questionnaire | Group | Mean confidence level<br>(score of 1 to 5) | 95% Confidence interval |
| --- | --- | --- | --- |
| Pre-intervention | Control (Group B) | 2.3 | [2.0; 2.6] |
|  | Experimental (Group A) | 2.1 | [1.8; 2.4] |
| Post-intervention | Control (Group B) | 3.2 | [2.9; 3.5] |
|  | Experimental (Group A) | 3.2 | [2.9; 3.5] |

**Figure 4:**
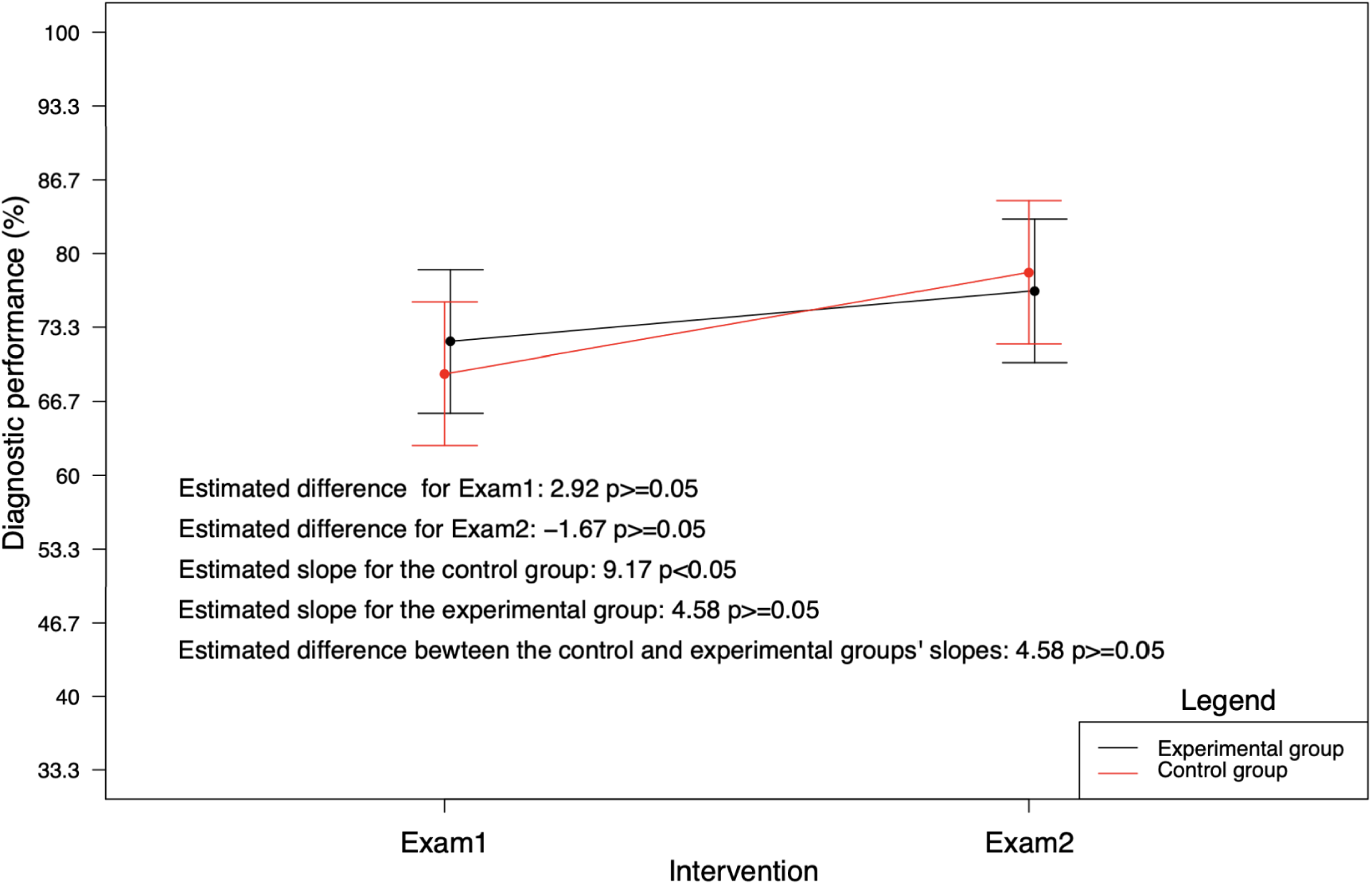
Participant’s diagnostic performance for the practice test (Exam1) and the final test (Exam2)

**Figure 5:**
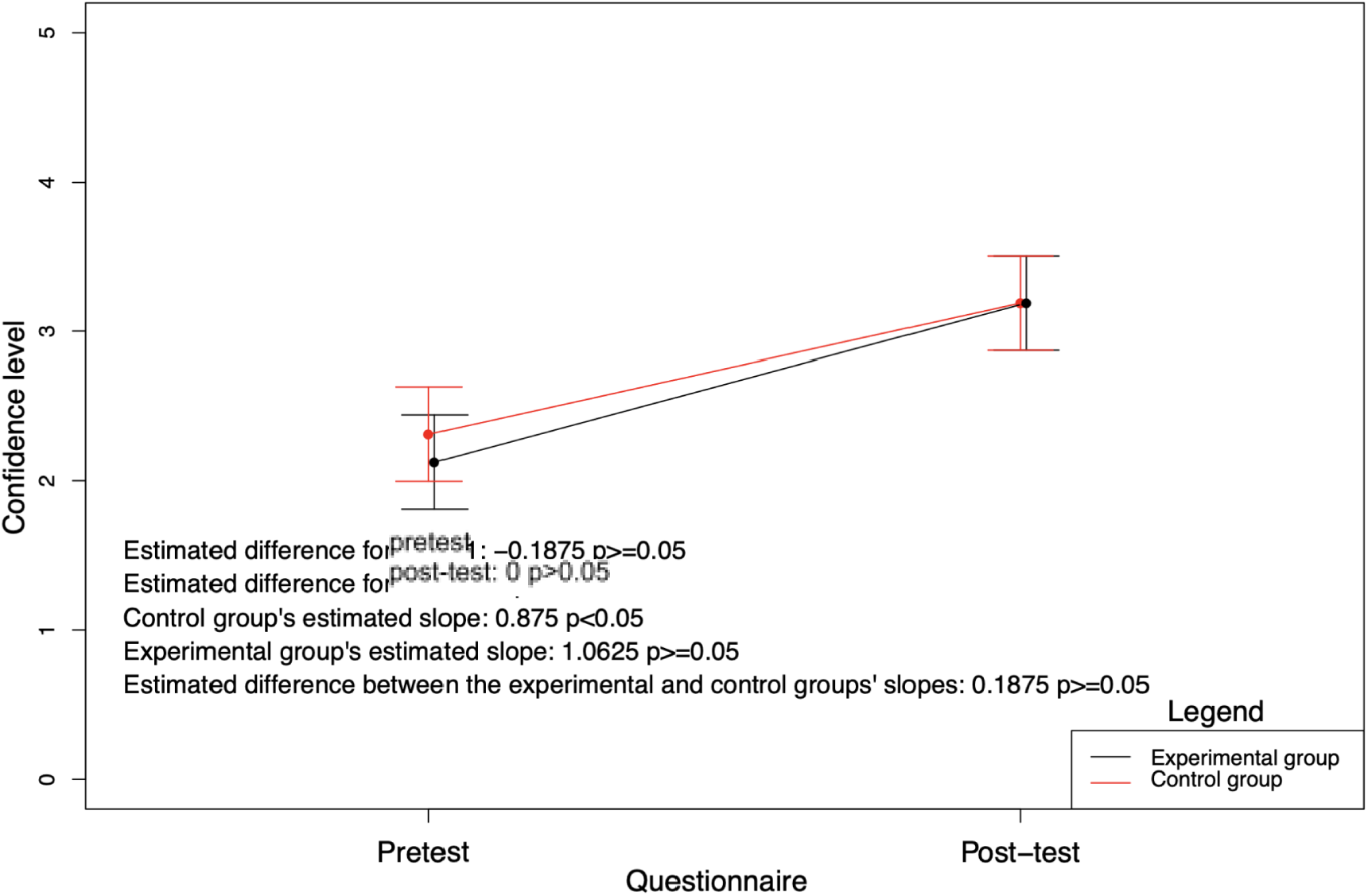
Participants’ confidence level in the pre-intervention and post-intervention questionnaires

### 3.1 Pre-intervention survey results

The analysis of the pre-intervention questionnaire results revealed no statistical difference in the level of confidence in chest X-ray interpretation between both groups (p-value=0.41) (Table 2).

### 3.2 Exam1 and Exam2 results

Prior to the intervention, the control group’s mean diagnostic performance on Exam1 was 69.2% (95% CI [62.7 ; 75.7]) compared to 72.1% in the experimental group (95% CI [65.6 ; 78.6]). There was no statistically significant difference in diagnostic performance between the experimental and control groups in Exam1 (p-value=0.53). On Exam2 (post-intervention), the control group’s mean diagnostic performance was 78.3% (95% CI [71.85 ; 84.81]) compared to 76.67% in the experimental group (95% CI [70.19 ; 83.15]). These results were not statistically different (p-value=0.36). Student’s diagnostic performance for the practice test (Exam1) and the final test (Exam2) are summarized in Table 3. The MMRM estimates were plotted in Figure 4 via their least squares mean estimates for better visualization of each group’s diagnostic performance and their comparison. Chester’s impact on students’ diagnostic performance is graphically presented via least squares mean estimates along with their confidence intervals (Figure 4 and 5).

### 3.3 Post-intervention survey results

Post-intervention survey results are summarized in Table 3 and Table 4. On a scale of 1 to 5, five being the highest, the mean level of confidence in chest X-ray interpretation skills in the post-intervention survey was identical in both groups at 3.2 (95% CI [2.9, 3.5]). The mean variation in confidence level for students in the control group was 0.9 (95% CI [0.5-1.3]), while it was 1.1 (95% CI [0.6-1.5]) for students in the experimental groups. On a score of one to five, participants from the experimental group rated Chester’s usefulness at 3.69 (95% CI [3.39; 3.98]) and its ease of use at 4.25 (95% CI [3.79; 4.71]) and were 4.13 likely (95% CI [3.73; 4.52]) to use it in another studying setting. When asked about the way they used the AI tool, 93.3% reported that they formed their own answer first before verifying using Chester and 6.7% (1 participant) reported that they only used Chester when they were seriously doubting their own answer.

**Table 4:** Participants’ answers to the post-intervention questionnaire

| Group | Question | Mean value (95% confidence interval) |
| --- | --- | --- |
| Experimental<br>(Group A) | For having used Chester during your study period (Exam1), how useful do you consider this AI tool to be? (1 = very harmful, 5 = very useful) | 3.69 (3.39; 3.98) |
|  | How easy do you consider this AI tool to be to use? (1 = very difficult, 5 = very easy) | 4.25 (3.79; 4.71) |
|  | How willing would you be to use this AI tool to study or learn the interpretation of chest X-rays in another setting? (1 = not at all, 5 = very ready) | 4.13 (3.73; 4.52) |
|  | How much do you trust this AI tool? (1 = not at all, 5 = complete trust) | 3.44 (3.04; 3.84) |
|  | How strongly do you recommend Chester to other medical students for learning or studying chest X-rays? (1 = not at all, 5 = strongly) | 4.00 (3.75; 4.25) |
|  | During your study period, how much did you use Chester compared to other available resources? (1 = only Chester, 5 = only other resources) | 2.94 (2.26; 3.62) |
|  | For having used Chester during your study period (Exam1), to what extent do you consider yourself to have been advantaged or disadvantaged compared to your peers in the other group on the final exam (Exam2 )? (1 = very disadvantaged, 5 = very advantaged) | 3.88 (3.48; 4.27) |
| Control<br>(Group B) | For not having used Chester during your study period, how useful do you think the tool was to your peers in the other group? (1 = very harmful, 5 = very useful) | 3.69 (3.22; 4.15) |
|  | How willing would you be to use an AI tool to study or learn the interpretation of chest x-rays in another setting? (1 = not at all, 5 = very ready) | 4.19 (3.78; 4.60) |
|  | For not having used Chester during your study period (Exam1), to what extent do you consider yourself to have been advantaged or disadvantaged compared to your peers in the other group during the final exam (Exam2)? (1 = very disadvantaged, 5 = very advantaged) | 2.38 (2.07; 2.68) |

Regarding their satisfaction with the experiment, 90.0% of total participants answered “yes”, “liked”, “appreciated”, “satisfied”, “useful” or “excellent experiment” and 10% declined to answer. Further information can be found in Table 4. Overall, students shared highly positive comments regarding the experiment and the tool. A few examples include: “practical tool when testing one’s ability to read radiographies no matter where we are”; “could be useful to help students learning”; “remarkable ease of use”; and “appreciated the experiment, it allowed me to practice my chest X-ray interpretation skills” (translated from French). Out of all the students who were offered the option to use Chester, only one participant did not use Chester at all, explaining that the tool was not intuitive enough to them.

## 4. Discussion

Given the novelty of the technology, there has been, to this day, only two other studies testing the impact of an AI tool in the radiological education of medical trainees, namely an American trial testing a brain MRI analysis tool (ARIES) on the diagnostic performance of four radiology residents (13) as well as a randomized controlled trial conducted by a Taiwanese team on 34 fifth-year medical students (20). In the latter study, researchers used a program called HipGuide to determine the presence or absence of hip fracture on pelvic X-rays (PXRs); the software also included a saliency map feature (20). Participants were randomized on a 1:1 basis and assigned to either an experimental group or a control group (20). Both groups took a prelearning test (100 images) and a post-learning test (100 images) (20). Participants in the experimental group took a third test between the prelearning and post-learning tests consisting of 100 AI-augmented PXRs with saliency maps shown by HipGuide (20).

While the Taiwanese study showed significant difference (p<0.01) between the experimental group’s prelearning score (75.73±10.58) and post-learning score (88.87±5.51), but no statistical significance (p=0.264) between the control group’s prelearning score (75.86±11.36) and post-learning score (78.66±14.53), the findings were reversed in this study with a statistically significant improvement in the control group’s score between Exam1 and Exam2 (slope: 9.17, p<0.05), but no statistically significant improvement in the experimental group’s score between Exam1 and Exam2 (slope: 4.58, p>0.05). This raises the question of whether using an AI tool (in this case, Chester), especially one with limited accuracy, could potentially stall participants’ acquisition of image interpretation skills and even hinder it.

This being said, many reasons could explain the discordance in results between the two studies. Firstly, HipGuide was a more performant algorithm than Chester with a performance of 91% on the test set, compared with Chester’s 60% accuracy. However, one could argue that HipGuide and Chester were fundamentally different programs, each charged with a specific task: while HipGuide detected the presence or absence of hip fractures, Chester identified 14 different radiological findings with quantified levels of probability. Secondly, both studies used a small number of participants (the Taiwanese study had 30 participants after accounting for attrition, while this study had 32 participants), which could have resulted in type II errors when it comes to the lack of statistically significant improvement in the results of this study’s experimental group and/or the Taiwanese study’s control group between the first test and the second test. Thirdly, many confounding biases could be found in the Taiwanese study. For example, the authors only employed 1:1 randomization and did not control for prior experience in medical image interpretation. Furthermore, participants in the experimental group were given 100 more images to improve their skills (using HipGuide) between the pre-learning test and the post-learning test, while participants in the control group only completed a pre-learning test and a post-learning test. Consequently, the difference in improvement between the experimental group and the control group could be attributed to the fact that participants benefited from 100 more examples to perfect their diagnostic skills, regardless if they used an AI tool or not. These confounding factors may have resulted in type I errors when it comes to the improvement of their experimental group’s results between the first test and the second test.

Compared to the Taiwanese study, our study is novel in the way that it examines the qualitative aspects of introducing an AI radiology assistant in medical education, namely its impact on participants’ level of confidence and their subjective appreciation of the tool.

Regarding participants’ level of confidence, the authors believed that novelty bias and the fact that students in the experimental group knew they had access to a supplementary tool would bolster their confidence level. While the mean variation in confidence level for students in the experimental group (slope: 1.0625) was greater than the one of the control group (slope: 0.875), the experimental group’s slope was not statistically significant (p>0.05), which disproved the initial hypothesis. Beyond a type II error, the lack of statistically significant improvement in confidence scores could be explained by the fact that all students received feedback after completing Exam1 and Exam2. Given that there was no statistically significant difference between the two groups’ results in Exam2, it comes as no surprise that post-test confidence levels did not show any substantial change in the experimental group.

Regarding participants’ appreciation of the experiment and the tool, due (once again) to novelty bias as well as the tool’s intuitive interface, the authors expected high scores in participants’ appreciation of the tool’s ease of use and the perceived advantage it granted them, as well as their use of it relative to other available resources. This hypothesis was corroborated by the results. What surprised the researchers, however, was participants’ perceived usefulness of Chester, their trust in it, their willingness to use it in another setting, and how much they recommended it to colleagues, considering that participants were warned of the tool’s limited performance prior to the intervention and had access to feedback after each test. This could be indicative of novelty bias and participants’ lack of confidence in their own skills as undergraduate medical students. If these results generalized to physicians, they could be cause for concern: would doctors, too, tend to trust an AI tool over their own clinical judgment, regardless of the algorithm’s performance? Finally, participants from both groups overwhelmingly showed appreciation for the experiment, which was anticipated by the authors, as undergraduate medical students’ exposure to radiological education and opportunities to practice their skills remain minimal (21,22).

Over the last decade, radiology has grown to uphold a more substantial role in medical education and previous research advocates for integration of radiologic education into the medical curriculum (21–28). However, radiologic education is still lacking. Therefore, we designed a pilot randomized study to fill the gap in the literature regarding AI tools and the feasibility of its use in the medical curriculum as well as to explore the acceptability of such a tool among medical students. This investigation is focused on improving education, specifically through the augmentation of existing educational methods such as textbooks and annotated radiology images (29–31) using an AI-based radiology assistant.

While the principle of Chester does not differ from annotated X-ray images, the advantage of using an AI tool over annotated X-ray images is that it can be used easily and quickly on any frontal chest radiographs the student encounters. In this study, Chester is strictly used to analyze images; students have to integrate the system’s input with other clinical information given in the question. Therefore, critical and clinical thinking skills remain paramount. In our research, Chester performed with 60% accuracy on Exam1. Thus, a participant in the experimental group who would have blindly trusted Chester’s prediction for each image would have only scored 9/15 in Exam1. In other words, any participant who scored above 60% would have had to rely on more than the tool itself.

Although the authors would have preferred to use a tool with a higher performance on “real-life” data, a concept in computer science called Out-of-Distribution (OOD) generalization, this issue commonly plagues machine learning models (32). Interestingly, our results show that the majority of participating students enjoyed using an AI tool in an education setting even if its diagnostic performance was merely 60%.

A key objective of this pilot study was to assess students’ use of a tool which they know is imperfect, something future physicians would have to contend with if AI algorithms are one day deployed in clinical settings. Indeed, it is the authors’ opinion that such as no predictive algorithm is 100% accurate, no tool used in medicine nowadays can be trusted entirely. Because no piece of the diagnostic puzzle, whether it be the patient’s history, the laboratory results, or medical imaging reports, presents the full picture, it is the clinician’s duty to interpret and, in some cases, disregard the data provided by those pieces. In this context, AI tools can help in the diagnostic puzzle but they do not necessarily give the full picture and should therefore be regarded by the clinician with a critical eye as simply as another tool instead of being welcomed as the medical gospel.

Some strengths of the study include its robust methodology, its use of stratified randomization, its lack of attrition, and the investigators’ efforts to isolate the intervention (the use of an AI radiology assistant) with a two-arm trial, all of which assured the study’s internal and external validity. As noted earlier, these strengths distinguish the study from previous work testing AI tools in radiological education settings.

Despite our strengths, our study has several noteworthy limitations. First, as an exploratory pilot study, we had a small sample size. This inevitably limits the external validity and the statistical power of our research. Second, Chester had a poor diagnostic performance of only 60% in Exam1 even if the cases selected seemed adequate for a third-year medical student level. While radiologists are exposed to many biases already (33), AI tools are also subject to their own unique limitations (34). Due to Chester being trained on labelled images, its performance is naturally affected by labelling biases: discrepancy between the radiologist’s, clinician’s, and automatic labeller’s understanding of a radiology report (35); errors in labelling due in part to automatic labellers (36,37); framing bias or limitations in objectivity (33,38,39); and interobserver variability, (40) to only name a few. Moreover, one limitation of the AI radiology assistant tool used in this experiment is that it only processes frontal X-ray images, which could potentially affect the performance and learning process of students.

Several reasons can explain Chester’s poor performance. AI systems almost always suffer from the black box problem (41), meaning that, as efficient as the algorithms can be, they do not grant us access to their inner workings. This setback can have devastating consequences in a field like medicine. To combat this issue, Chester’s developers implemented a saliency map for findings scoring high enough in probability. This allows users to visualize the pixels which the program draws from in order to predict a particular finding. This feature is, however, not necessarily indicative of the location of the finding. For example, the saliency map for the finding of “consolidation’ could highlight a region with no consolidation, but rather pixels in the spine. Far from a sign that the system is wrong, this could mean that the algorithm used those pixels to determine the position of the lungs, where the consolidation truly lay. Finally, the saliency map feature should not distract the user from the fact that Chester does not truly “understand” what a lung, a heart, or pneumonia truly are; rather, it processes pixels and recognizes patterns based on the shade and relative position of those pixels.

Most importantly, our study highlights the importance of testing a newly developed educational tool before using it at a larger scale. While the results of the pilot study are inconclusive for the moment, they demonstrate proof of concept for a repeat experiment with a larger sample. More importantly, this study establishes a robust methodology to evaluate AI tools in radiological education, regardless of their performance. Finally, the appreciation shown by participants both for the tool and the experiment itself is indicative of a large and, to this day, untapped potential with respect to AI applications in medical education. In the context of the COVID-19 pandemic, which has likely affected an entire generation of medical students and graduates (37,38), as well as the growing trend of remote learning, we believe that further research should be conducted on the applications of AI and development of AI tools in medical education.

## Data Availability

All data produced in the present study are available upon reasonable request to the authors.

## APPENDIX – FIGURES AND TABLES

**Flowchart 1:**
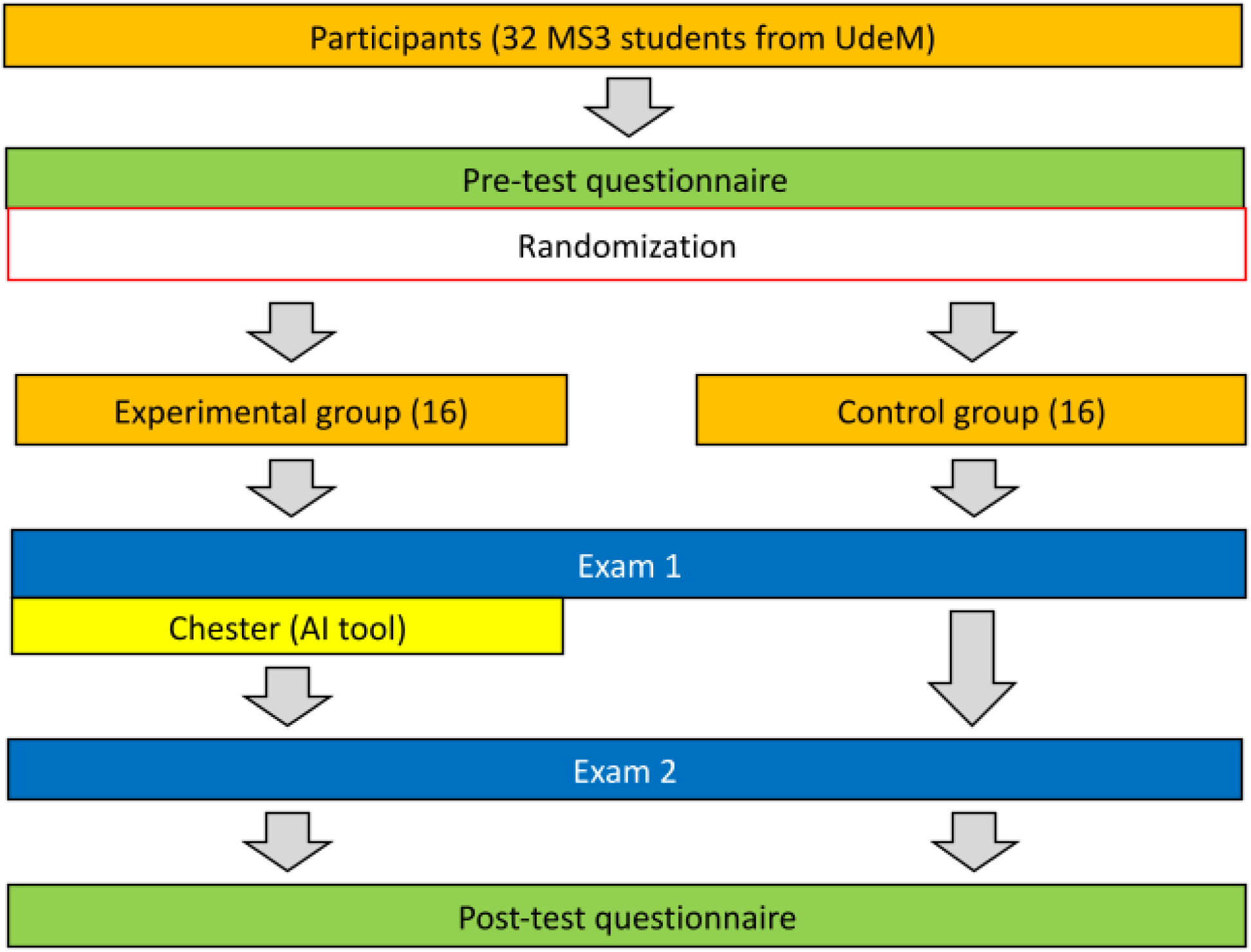
Overview of the intervention, including the pretest and post-test questionnaire, randomization process, Exam1, and Exam2

## Footnotes

1 https://mlmed.org/tools/xray/

2 https://youtu.be/yP4EAZ2E6-s

3 https://radiopaedia.org/

## Notes

<u>Financial Disclosure Statement:</u> No conflict of interest to declare. No funding was received for this article.

### Competing Interest Statement

The authors have declared no competing interest.

### Funding Statement

This study did not receive any funding.

### Author Declarations

The Institutional Review Board of the University of Montreal gave ethical approval for this work

### Summary of Updates

Changed email of the first author.

